# Spatial Patterns of Dengue Incidence in Nepal During Record Outbreaks in 2022 and 2023: Implications for Public Health Interventions

**DOI:** 10.1101/2024.11.06.24316870

**Authors:** Simrik Bhandari, Jason K. Blackburn, Sadie J. Ryan

## Abstract

Dengue, first reported as a travel case in Nepal in 2004, was initially confined to the lower plains, but has spread to higher elevations. It occurred as large outbreaks in 2022 and 2023 (54,784 and 51,243 cases, respectively), reaching every district (n=77). We calculated dengue incidence in each district in 2022 and 2023, via digitizing case data from Nepal’s Ministry of Health and Population, and 2021 census data from the National Statistics Office. Incidence and peak dengue months in each year were mapped, and spatial clusters (hotspots and cold spots) and spatial outliers of incidence rates for the two years were identified using Local Moran’s I. In 2022, district-wise peak cases occurred in August – October. One hotspot (High-High) including six districts around Kathmandu, and one cold spot (Low-Low) comprising eight high elevation districts in Nepal’s northwest region were identified. In 2023, cases peaked March-November, indicating more distributed peaks, starting earlier; hotspots shifted to north-central and eastern regions, and a Low-High outlier district in the central region was identified. Identifying timing of peaks, and spatial clusters of dengue incidence can inform targeted management, improving effectiveness and cost-efficiency. This study provides a baseline examination of recent dengue in Nepal, highlighting timing and spatial clustering in incidence. The mountainous northwest cold spots align with expectations of fewer mosquitoes due to geography and climate. However, dengue peaked in all 77 districts over three months in 2022, suggesting ecological and climatic factors may no longer be effective barriers.

## Introduction

Dengue fever, a mosquito-borne viral infection, is a significant global public health problem, leading to an estimated 390 million infections annually ^1^. Dengue virus (DENV) is an arbovirus in the family Flaviviridae, transmitted by *Aedes* spp. mosquitoes, primarily *Aedes aegypti* and *Aedes albopictus* ^2^. DENV infection can present when a range of symptoms. Infected individuals may be asymptomatic, experience mild fever, or suffer from severe forms of the disease, such as dengue hemorrhagic fever (DHF) and dengue shock syndrome (DSS). DHF is characterized by high fever, severe headache, bleeding (such as nosebleeds or gum bleeding), and abdominal pain. DSS, a more severe progression, includes symptoms like a rapid, weak pulse, cold, clammy skin, and low blood pressure, leading to potentially life-threatening shock. Dengue is also known as ‘bone-breaking fever’ due to the severe muscle and bone pain it can cause ^3^. The disease is more prevalent in warmer regions near the equator, particularly in tropical and subtropical areas ^4,5^. The dengue virus has four distinct serotypes (DENV-1, DENV-2, DENV-3, and DENV-4). Infection with one serotype provides immunity to that specific serotype but increases the risk of severe disease if a person is subsequently infected with a different serotype ^6^. This phenomenon complicates the development of an effective dengue vaccine.

Dengue impacts livelihoods, and the burden affects healthcare systems, especially in regions with tropical and subtropical climates ^4,5^. In Nepal, dengue fever was first reported in 2004, and has evolved into a significant public health issue, now considered endemic in the country. All four dengue virus serotypes are now present and co-circulating in Nepal, and both *Aedes albopictus* and *Ae. aegypti* have been identified as the key transmission vectors in Nepal ^7^.

Initially confined to the warmer southern lowland Terai region of Nepal, dengue has spread to higher elevations and previously unaffected areas ^8,9^. Climate change has been shown to have influenced mosquito breeding patterns and expanded *Aedes* spp. mosquito habitats in Nepal ^10,11^. Additionally, excessive rainfall, poor waste management, and rapid unplanned urbanization create ideal breeding grounds for these mosquitoes. Urbanization supports mosquito breeding habitat by creating diverse artificial aquatic habitats, such as buckets and flowerpots, which provide ideal conditions for *Aedes* spp. ^12,13^; these processes have been underway in Nepal, as in much of the world, over the past two decades. The open border with India also plays a role in dengue transmission in Nepal, facilitating cross-border introductions and transmission of the disease ^14^.

Nepal’s health system faces considerable challenges in controlling dengue. These include difficulties in monitoring, early detection, and diagnosis. Significant outbreaks in 2010, 2013, 2016, 2019, 2022, and 2023 highlighted these challenges ^15^. In 2020 and 2021 there was a decrease in reported cases, likely due to lockdowns and restricted movement, but underreporting due to a focus on COVID-19 may have played a role ^16^. The year 2022 saw a significant rise in dengue in Nepal, with an alarming 54,784 cases recorded ^17^. In 2023, although there was a decrease in the reported cases, the number was still high with 51,243. Both years saw the cocirculation of DENV-1, DENV-2, and DENV-3 ^18,19^. In response to increasing case numbers and recent record years, the government has taken steps to mitigate outbreaks by offering free testing and treatment for dengue and focusing on prevention and control measures ^20^.

Identifying where regions are experiencing higher or lower than expected disease incidence can inform targeted public health interventions like mosquito control programs and public health education, ensuring efficient resource allocation in high-risk zones. Geospatial methods are crucial to studying disease incidence and spread, particularly in a management context ^21,22^. The Local Indicators of Spatial Association (LISA) statistic is often used in exploratory spatial data analysis (ESDA) frameworks to quantify spatial clustering or dispersion of disease cases or incidence in a study area, including studies of dengue and other arboviral diseases in emerging and endemic settings ^23,24^. Identifying spatially discrete areas of significantly high (e.g. hotspots) or low (e.g. cold spots) disease activity within administrative functional boundaries, can frame responses to public health events. In this study, we mapped district level dengue incidence in Nepal in 2022 and 2023, identify peak case months by district, and identify patterns of spatial heterogeneity in incidence across the country, using LISA methods.

## Methods

### Study Area

Nepal is a small (147,516 km^2^) mountainous, landlocked country in South Asia, surrounded by China to the north and India to the south, east, and west. The population in 2021 was 29,164,578, with a population growth rate of 0.92% ^25^. The country exhibits diverse topography and ecology with low-lying Terai plains in the south, hilly midlands, and the high Himalayas mountains in the north ^26^ (Figure 1). Nepal experiences four distinct climatic periods during the year: the pre-monsoon season (March to May), the monsoon season (June to September), the post-monsoon period (October to November), and the winter months (December to February) ^27^. Administratively, Nepal is currently divided into seven provinces and 77 districts.

**Figure 1.**
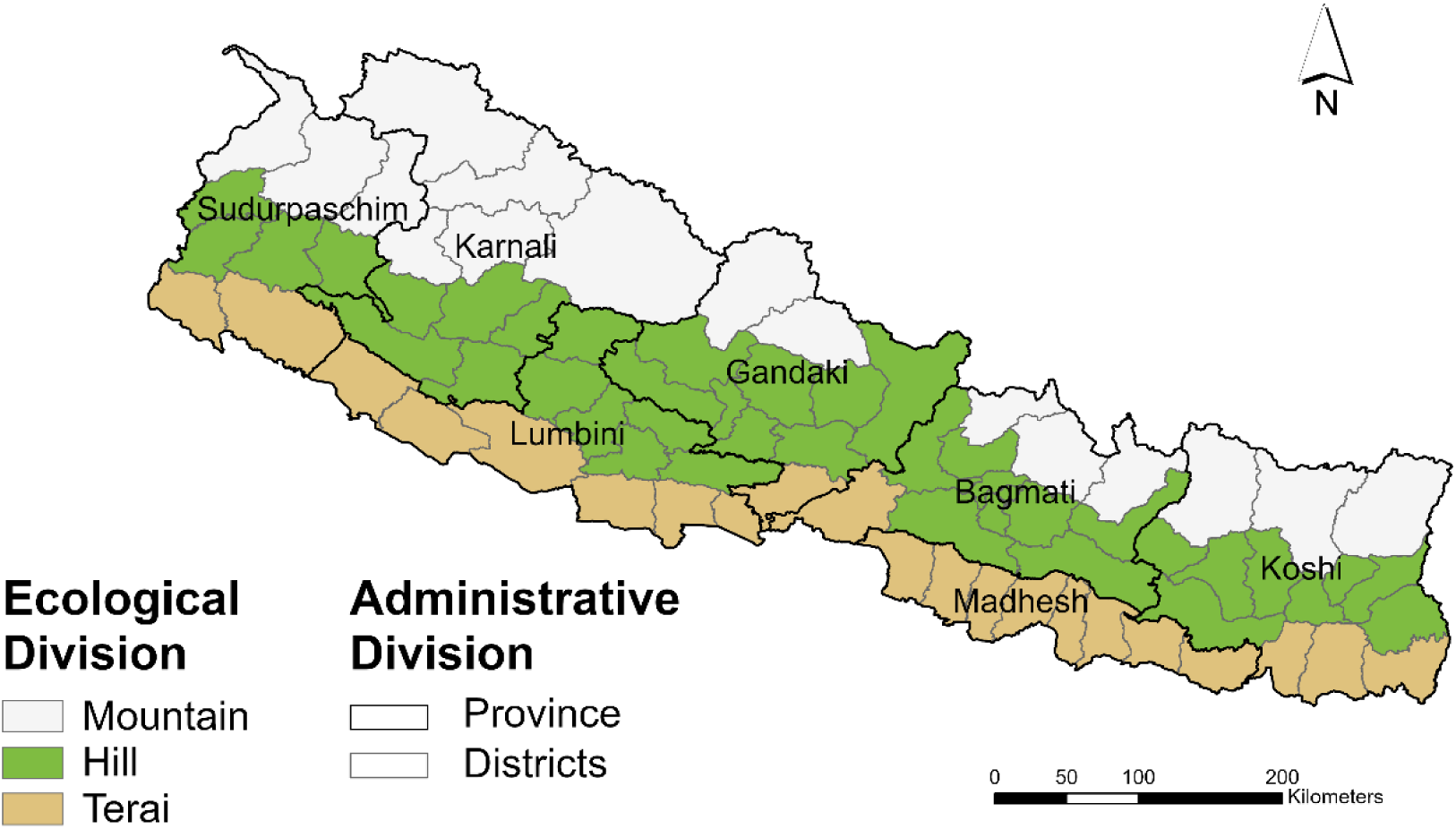
Ecological and Administrative Divisions of Nepal. The 77 districts and 7 provinces of Nepal, overlaid on three ecological divisions: Mountain (white), Hill (green), and Terai (brown).

### Data Acquisition and Processing

Monthly district-level dengue cases reported for 2022 and 2023 were obtained from the Epidemiology and Disease Control Division (EDCD) of the Ministry of Health and Population, Nepal ^28,29^. Population data for each district was obtained from the National Census Report 2011^30^ and 2021^25^. Administrative division shapefiles (geographic coordinate system WGS 1984) were downloaded from the National Geo Portal (https://nationalgeoportal.gov.np/). Dengue cases and population datasets were digitized and geospatially joined to district polygons for analyses and visualization using ArcGIS Pro^31^.

#### Population projections

District level population estimates for 2022 and 2023 were projected from 2011 and 2021 census data using an exponential growth model ^32,33^. The annualized growth rate (r) was calculated as:

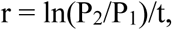

where P_1_ is the 2011 population count, P_2_ is the 2021 population count, and t is the number of years between the two censuses.

This growth rate was then used to project the population for the required years:

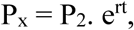

where P_x_ is the population estimate for the target year x, and t is years from the 2021 census to the target year (2022, 2023).

#### Case Timing Peaks

The months of peak case number, by district, were recorded, for 2022 and 2023, and geospatially joined to district polygons for mapping.

#### Incidence rates

Annual dengue incidence rates per 100,000 were calculated using GeoDa 1.20 ^34^. Dengue incidence was calculated as the total number of dengue cases in a reporting year in each district divided by its total population.

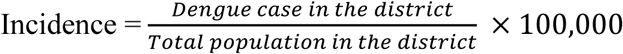

#### Spatial Clustering

Global Moran’s I provides an indicator of spatial clustering, to assess whether a phenomenon is spatially random, clustered, or dispersed on a landscape. This index takes values −1 – 1, from dispersed to clustered, and through simulations, permutations can provide a distribution of values, such that a statistic of significance can be assessed. This was applied to district-wise incidence in each 2022 and 2023, and 999 permutations run in GeoDa software to derive z-scores and assess significance at 0.025, to account for two years in this analysis.

#### LISA Analysis

Local Moran’s I statistic is a popular tool used in spatial analysis to identify disease patterns ^35,36^. It analyzes whether similar or dissimilar values in a dataset are geographically clustered, to detect areas of hotspots, cold spots, and outliers^37^. When working with small population areas, rate data, such as incidence, can be subject to instability due to small populations in sub-regions. Smoothing techniques in spatial analysis are applied to adjust rates to account for this instability ^38^. To assess any potential rate instability in this study, both Empirical Bayes Smoothing (EBS) and Spatial Bayes Smoothing (SBS) were applied to the incidence rates and compared to raw rates using box plots (Figure S1).

Local Moran’s I was calculated for the raw incidence rate using a queen contiguity spatial weight matrix with 999 permutations at a 0.025 significance level (to account for analyses spanning two years), identifying significant clusters and outliers. Maps of significant hotspots and cold spots for each year were constructed to visually assess spatial patterns.

## Results

Dengue was first reported in Nepal in 2004, as a travel case, and was initially constrained to the lowland Terai region. Until 2010, reported cases remained fewer than 50 each year; in the next decade, annual cases had increased to being reported in the several hundred, with 2016 and 2017 first seeing numbers exceeding a thousand. In 2019, an outbreak of nearly 18,000 reported cases brought attention to this public health threat, and its expansion in the country. In 2020 and 2021, perhaps due to redirected resources for the COVID-19 pandemic, reported cases were in the hundreds. In 2022 and 2023, echoing dengue outbreaks around the world, Nepal had outbreaks with case numbers reported more than double that of any prior year on record (Figure 2).

**Figure 2.**
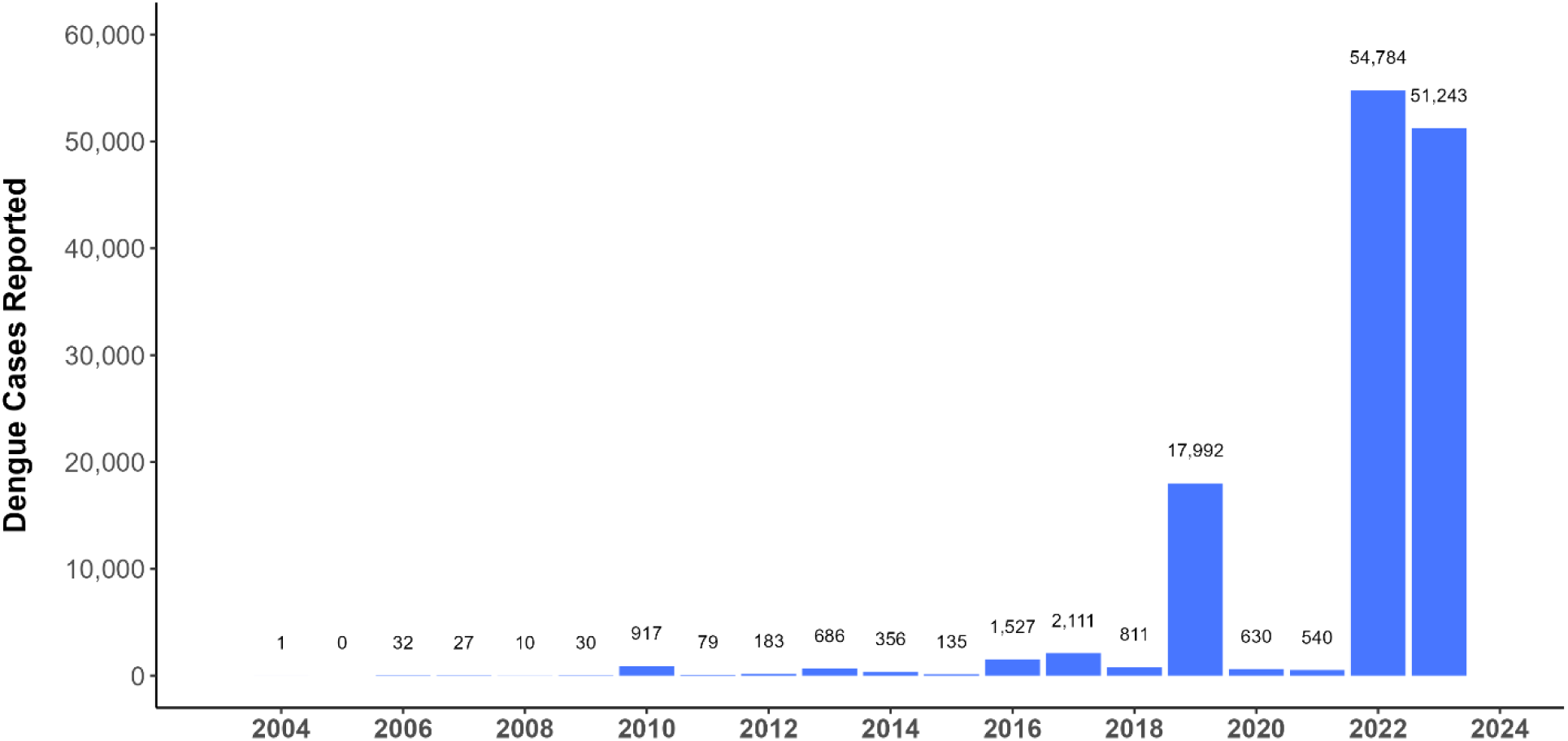
Yearly reported dengue cases in Nepal.

In 2022, in Nepal, there was a total of 54,784 reported dengue cases, and district level incidence ranged from a low of 4.21 to a high of 1,716.89. In 2023, the total number of reported cases was 51,243, with a district level incidence range of 2.49 to 2,242.44. The districts with the highest incidence rates were not identical between the two years, and in 2023, high incidence rates occurred further east than in 2022 (Figure 3).

**Figure 3.**
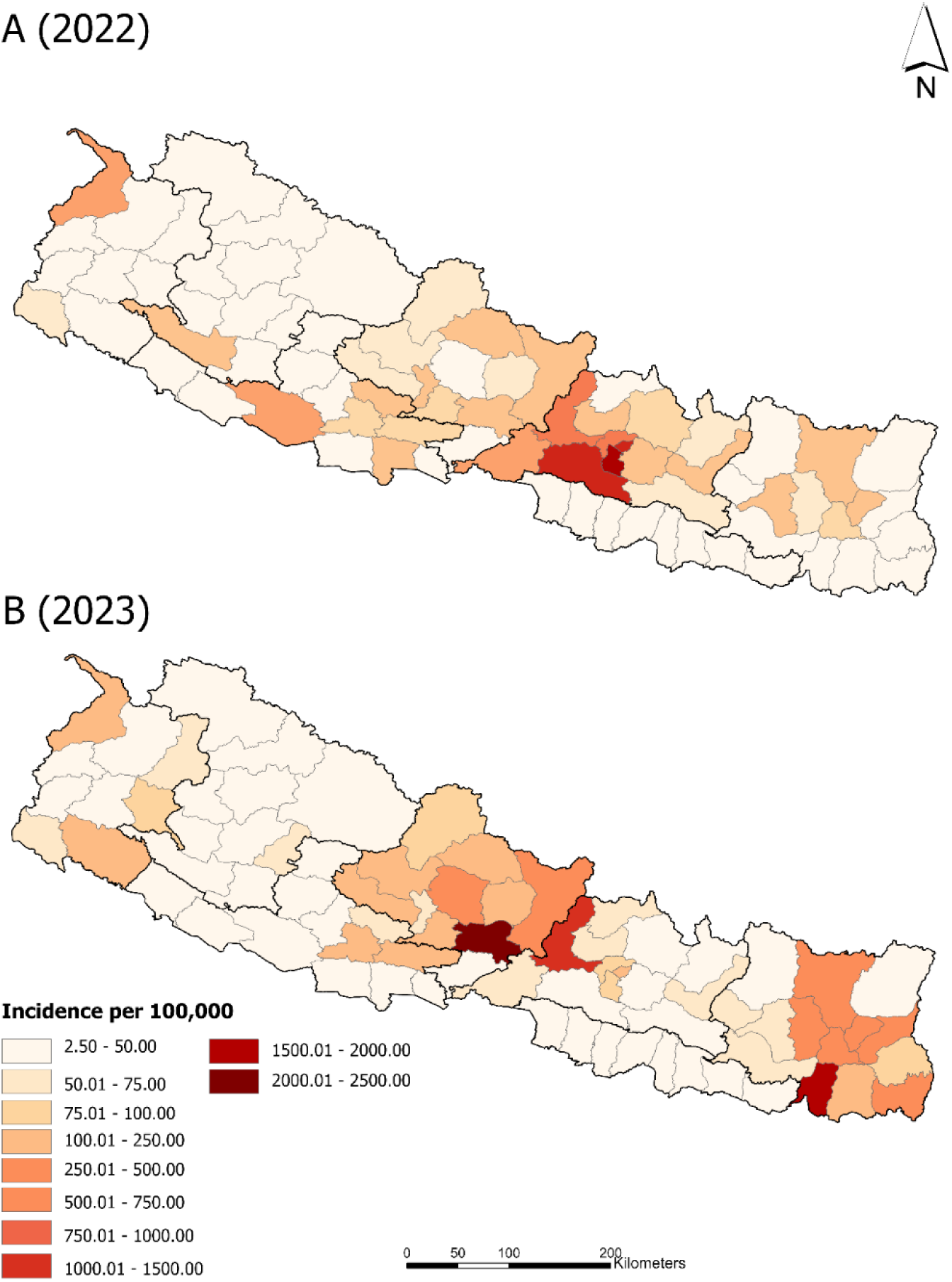
Incidence per 100,000 by districts of Nepal in 2022 and 2023.

The district level peak case month map for 2022 year illustrates that most dengue cases were reported in the months of August, September, and October (Figure 4A), with no obvious spatial patterns, suggesting that the whole country experienced one epidemic peak season. In 2023, cases were at their highest in some districts as early as March and peaked as late as November in the mountainous region (Figure 4B). The pattern of peak months in 2023 was more heterogeneous, although this was only visually assessed.

**Figure 4.**
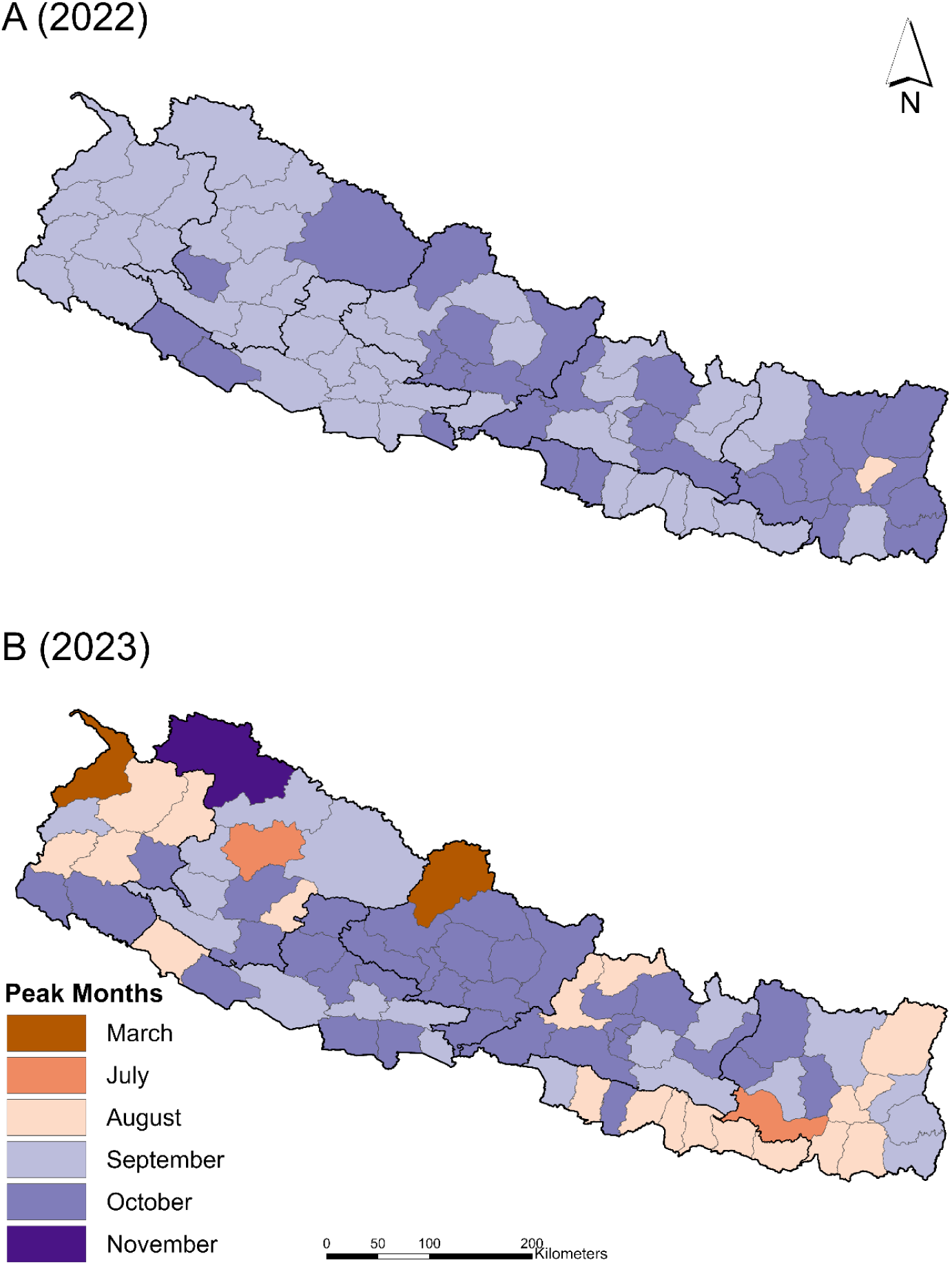
Month of peak number of dengue cases by district in 2022 and 2023.

Global Moran’s *I* indicated clustering in both 2022 and 2023, with positive values of *I* (Table 1). Based on permutation tests, 2022 showed significant clustering at the global level, for our criterion of significance level (α=0.025).

**Table 1:** Global Moran’s *I* values for dengue incidence, aggregated to district for 2022 and 2023.

| Year | Total Dengue Cases | Moran's $I$ | Z-score | P-value |
| --- | --- | --- | --- | --- |
| 2022 | 54,784 | 0.472 | 7.4798 | **0.001** |
| 2023 | 51,243 | 0.095 | 1.7947 | 0.060 |

P-values in bold indicate statistically significant results. \*Moran's $I$ values range between -1 and 1, where negative values indicate dispersion and positive values indicate clustering

The LISA map for 2022 dengue incidence (Figure 5A) shows one hotspot and one regional cold spot. Six districts in the centrally located province of Bagmati (Kathmandu, Lalitpur, Bhaktapur, Kavrepalanchowk, Makwanpur and Dhading) had statistically higher dengue incidence and eight districts had statistically lower dengue incidence in the west (Humla, Mugu, Bajura, Kalikot, Jumla, Jajarkot, Rukum West, Surkhet).

**Figure 5.**
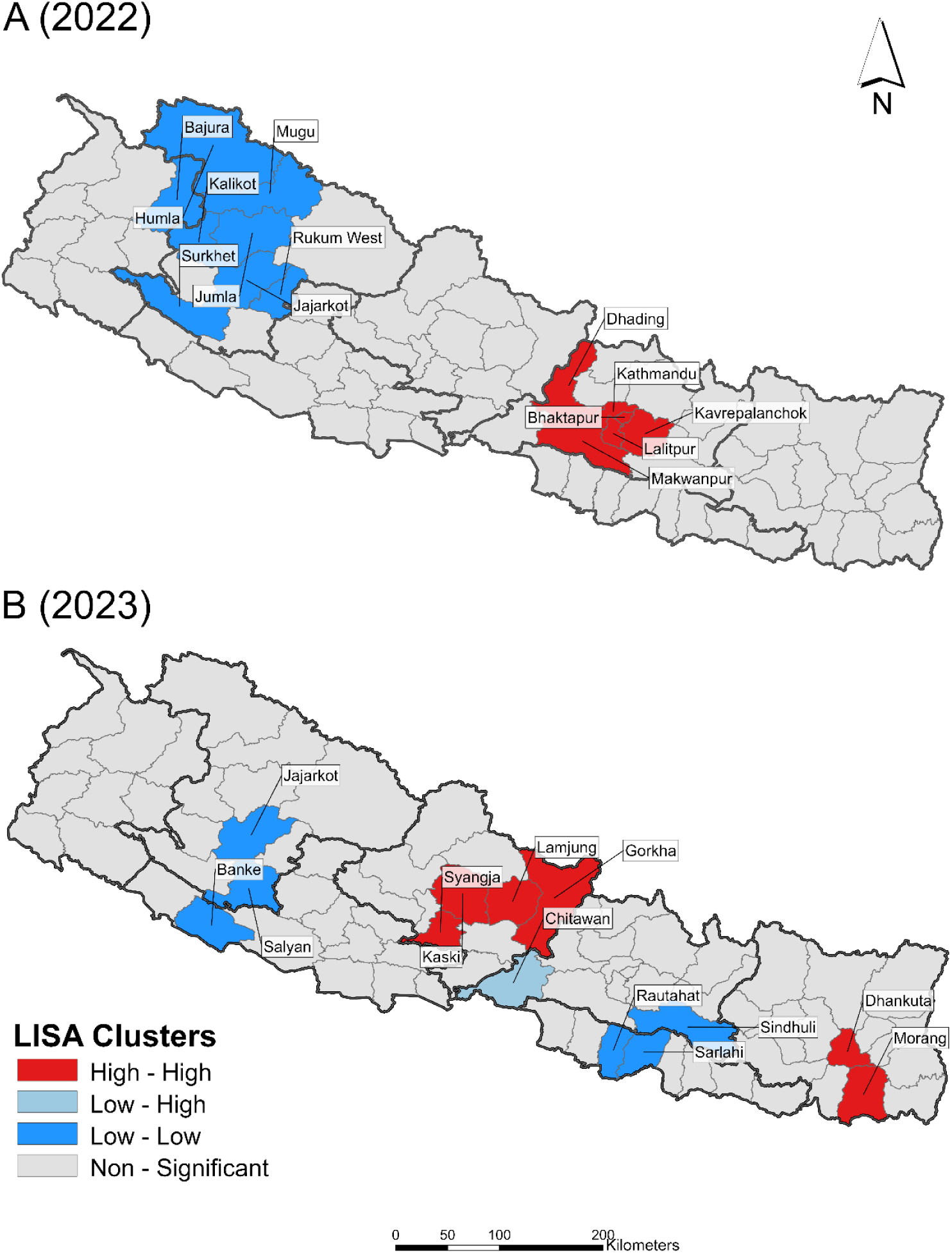
Hotspots and cold spots of dengue incidence as identified by Local Indicator of Spatial Association (LISA) for 2022 and 2023 in Nepal. Areas in red indicate clusters of districts where districts with high rates are surrounded by districts with similarly high rates, indicating a significant hotspot for dengue transmission. Blue areas represent low clusters, indicating districts with low rates surrounded by districts with similarly low rates. The area in light blue color indicates statistically significantly lower incidence than expected for its neighboring districts.

The LISA map for 2023 dengue incidence reveals a shift in clusters (Figure 5B) to two hotspots and two cold spots. Four centrally located districts (Gorkha, Lamjung, Kaski, and Syangjha) and 2 districts located to the east (Dhankuta, Morang) had statistically higher dengue incidence. Similarly, three districts in the west (Jajarkot, Salyan, Banke) and three centrally located districts (Sindhuli, Sarlahi, Rautahat) had statistically lower dengue incidence. One Low – High outlier in the Chitwan district also indicates a lower incidence than in neighboring districts.

## Discussion

In 2022 and 2023, Nepal experienced dengue outbreaks reporting more than double the number of cases of any prior year on record. In a country experiencing such rapid and expansive outbreaks of this emergent vector-borne disease, visualizing and understanding spatial patterns and timing can inform public health planning and intervention. Dengue cases were reported in every district of the country in 2022, which indicates that previously assumed climate and ecological barriers to establishment and transmission are no longer acting to curb its spread. Using exploratory geospatial approaches, we digitized and visualized the 2022 and 2023 dengue outbreak years for Nepal, based on published case data and census population projections.

Mapped peak case months by districts showed that in 2022, the highest case numbers occurred clumped in time, spanning August to October with no obvious heterogeneity in the country. However, in 2023, peak case numbers occurred in districts from March to November, with mountainous districts peaking both earliest and latest, and showing a patchier distribution of peak months across the country. Annual incidence maps showed that in 2022, very high incidence districts were largely clumped around Kathmandu, while in 2023, not only were these not the same districts showing higher incidence, but an emergence of higher reported incidence in eastern Nepal emerged. The distribution of incidence was clustered in both years at the global scale, but only significantly so in 2022. This was echoed in a shift in dengue hotspots and cold spots from the year 2022 to 2023. In 2022, the hotspot (high-high cluster of districts) identified around Kathmandu was also contained entirely within Bagmati province, while in 2023, the two identified hotspots were clusters of districts occurring in the two provinces to the west (Gandaki) and east (Koshi). Interestingly, one district in Bagmati became part of a cold spot in 2023, suggesting lower incidence than expected, and a second district in Bagmati (Chitwan) was a low-high outlier in 2023, indicating incidence rates significantly lower than expected, and not part of a cluster. Whether province-level shifts such as this could reflect the impact of population level immunity from the prior year at the district-level, or successful interventions, improved vector-control and prevention, requires follow-up studies leveraging approaches outside the scope of this study, but including serological surveillance, and cross-sectional or longitudinal health center level survey instruments.

It is important to note that this study relies on reported case data, and in a large and rapidly unfolding outbreak situation, the availability of diagnostic testing, accessibility to health facilities and implementation of intervention are unevenly distributed in space and time, which may create uneven data reporting, and over-representation of better resourced areas. While we might expect that areas with very sparse populations, where healthcare access is quite limited, such as the mountainous region, could lead to a lack of reporting, the presence of cases reported in every district in 2022 suggests that this did not lead to overly spatial biased outcomes in our study. However, we are cautious in our interpretation of the cold spot cluster of low-low incidence in the northwest mountainous region identified in 2022, as it may be more indicative of sparse reporting than existing prevention. While the sparse population in the mountainous districts could also distort incidence rate calculation, we found that the incidence rates were not unstable or impacted by population distribution for the purposes of conducting the ESDA analyses here.

Using local indicators of spatial association (LISA) methods, the study identified hotspots (i.e., districts with higher dengue incidence) indicating potential high-priority areas for targeted health measures in those regions. In the scenario of vector-borne disease management, these can indicate areas that may need increased intervention, such as spraying, larval source management (LSM), or increased allocation of hospital resources for palliative care and diagnostic testing. The shifts in identified hotspots between the two years in this study are worth further consideration, to explore how best to allocate limited resources. The study also highlighted cold spots, regions with lower than expected dengue incidence, which could result from how local health strategies, population immunity, or case reporting vary from place to place.

Nepal is a small country with diverse topography leading to a sharp climate and ecological gradients. It is experiencing rapid shifts in climate-induced impacts, which influences both human population distributions on the landscape, and the ecology of vector habitats. Understanding the dynamics underpinning the spatial patterns of a rapidly establishing vector borne disease described here through analysis of spatial human incidence data requires further investigation. Future research on the spatial distribution of dengue in Nepal should explore additional drivers (socioeconomic, climatic, and environmental influences) of dengue transmission.

## Supplemental Material

**Figure S1:**
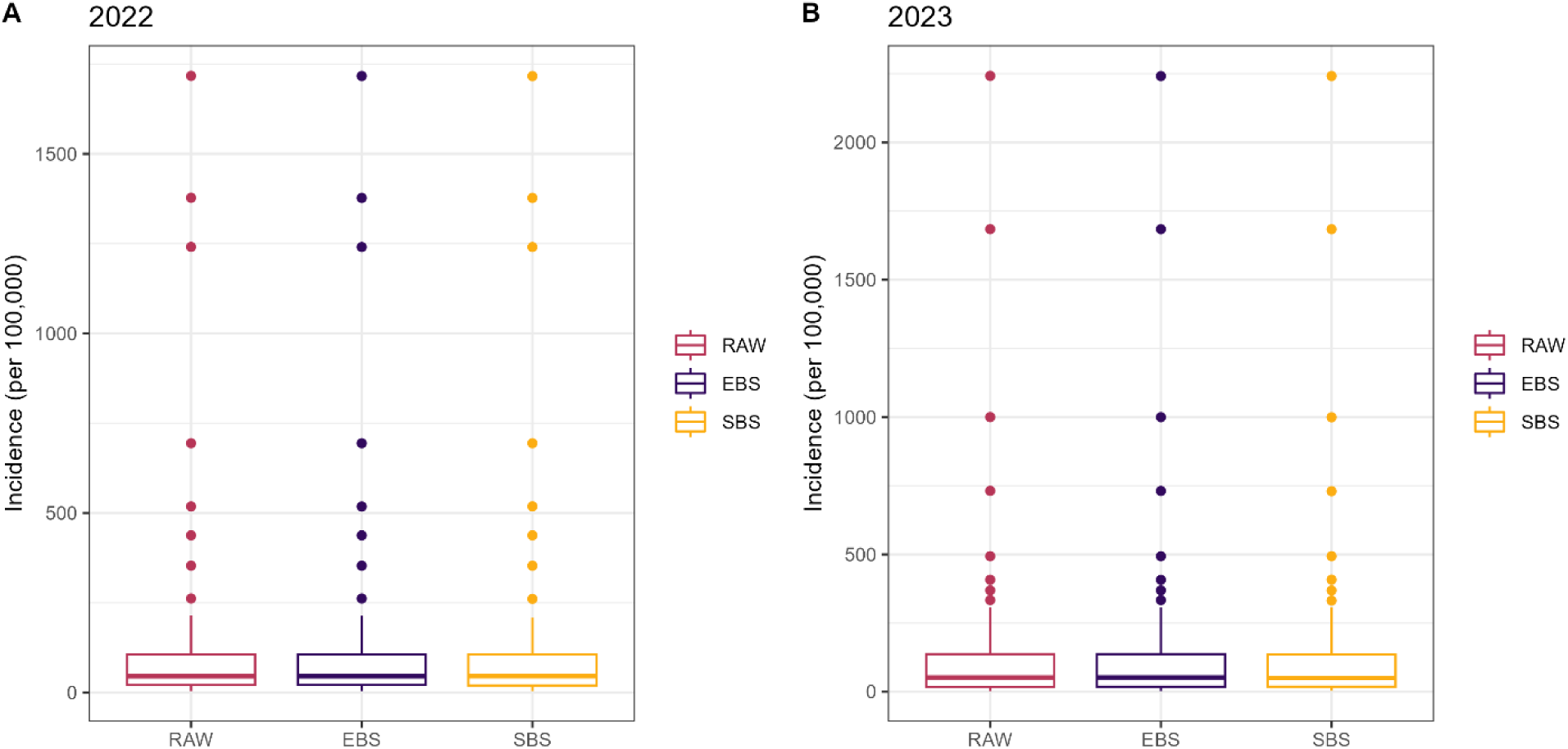
Boxplot comparison of incidence rates for A. 2022, and B. 2023, showing no appreciable differences among Unsmoothed (RAW), Empirical Bayes Smoothing (EBS), and Spatial Bayes Smoothing (SBS)

## Data Availability

All data produced in the present work are contained in the manuscript

## Disclosures regarding real or perceived conflicts of interest

The authors report no conflicts of interest.

## References

1. Bhatt S, Gething PW, Brady OJ, Messina JP, Farlow AW, Moyes CL, Drake JM, Brownstein JS, Hoen AG, Sankoh O, Myers MF, George DB, Jaenisch T, Wint GRW, Simmons CP, et al., 2013. The global distribution and burden of dengue. Nature 496: 504–507

2. De Almeida RR, Paim B, De Oliveira SA, Souza AS, Gomes ACP, Escuissato DL, Zanetti G, Marchiori E., 2017. Dengue Hemorrhagic Fever: A State-of-the-Art Review Focused in Pulmonary Involvement. Lung 195: 389–395

3. World Health Organization., 2009. Dengue guidelines for diagnosis, treatment, prevention and control : new edition

4. Gubler DJ., 2002. Epidemic dengue/dengue hemorrhagic fever as a public health, social and economic problem in the 21st century. Trends Microbiol 10: 100–103

5. Guzman MG, Halstead SB, Artsob H, Buchy P, Farrar J, Gubler DJ, Hunsperger E, Kroeger A, Margolis HS, Martínez E, Nathan MB, Pelegrino JL, Simmons C, Yoksan S, Peeling RW., 2010. Dengue: a continuing global threat. Nat Rev Microbiol 8: S7–S16

6. Gubler D., 1998. Dengue and Dengue Hemorrhagic Fever. Clin Microbiol Rev 11: 480–496

7. Kawada H, Futami K, Higa Y, Rai G, Suzuki T, Rai SK., 2020. Distribution and pyrethroid resistance status of Aedes aegypti and Aedes albopictus populations and possible phylogenetic reasons for the recent invasion of Aedes aegypti in Nepal. Parasit Vectors 13: 213

8. Dhimal M, Gautam I, Kreß A, Müller R, Kuch U., 2014. Spatio-Temporal Distribution of Dengue and Lymphatic Filariasis Vectors along an Altitudinal Transect in Central Nepal. PLoS Negl Trop Dis 8: e3035

9. Tuladhar R, Singh A, Varma A, Choudhary DK., 2019. Climatic factors influencing dengue incidence in an epidemic area of Nepal. BMC Res Notes 12: 131

10. Acharya BK, Cao C, Lakes T, Chen W, Naeem S., 2016. Spatiotemporal analysis of dengue fever in Nepal from 2010 to 2014. BMC Public Health 16: 849

11. Pandey BD, Pandey K, Neupane B, Shah Y, Adhikary KP, Gautam I, Hagge DA, Morita K., 2015. Persistent dengue emergence: the seven years surrounding the 2010 epidemic in Nepal. Trans R Soc Trop Med Hyg: trv087

12. Li Y, Kamara F, Zhou G, Puthiyakunnon S, Li C, Liu Y, Zhou Y, Yao L, Yan G, Chen X-G., 2014. Urbanization Increases Aedes albopictus Larval Habitats and Accelerates Mosquito Development and Survivorship. PLoS Negl Trop Dis 8: e3301

13. Wilke ABB, Chase C, Vasquez C, Carvajal A, Medina J, Petrie WD, Beier JC., 2019. Urbanization creates diverse aquatic habitats for immature mosquitoes in urban areas. Sci Rep 9: 15335

14. Pokharel P, Khanal S, Ghimire S, Pokhrel KM, Shrestha AB., 2023. Frequent outbreaks of dengue in Nepal – causes and solutions: a narrative review. Int J Surg Glob Health 6

15. Acharya KP, Chaulagain B, Acharya N, Shrestha K, Subramanya SH., 2020. Establishment and recent surge in spatio-temporal spread of dengue in Nepal. Emerg Microbes Infect 9: 676–679

16. Pandey BD, Ngwe Tun MM, Pandey K, Dumre SP, Bhandari P, Pyakurel UR, Pokhrel N, Dhimal M, Gyanwali P, Culleton R, Takamatsu Y, Costello A, Morita K., 2022. Has COVID-19 suppressed dengue transmission in Nepal? Epidemiol Infect 150: e196

17. Bijukchhe SM, Hill M, Adhikari B, Shrestha A, Shrestha S., 2023. Nepal’s worst dengue outbreak is a wake-up call for action. J Travel Med 30: taad112

18. Rimal S, Shrestha S, Pandey K, Nguyen TV, Bhandari P, Shah Y, Acharya D, Adhikari N, Rijal KR, Ghimire P, Takamatsu Y, Pandey BD, Fernandez S, Morita K, Ngwe Tun MM, et al., 2023. Co-Circulation of Dengue Virus Serotypes 1, 2, and 3 during the 2022 Dengue Outbreak in Nepal: A Cross-Sectional Study. Viruses 15: 507

19. Rimal S, Shrestha S, Paudel SW, Shah Y, Bhandari G, Pandey K, Kharbuja A, Kapandji M, Gautam I, Bhujel R, Takamatsu Y, Bhandari R, Klungthong C, Shrestha SK, Fernandez S, et al., 2024. Molecular and Entomological Characterization of 2023 Dengue Outbreak in Dhading District, Central Nepal. Viruses 16: 594

20. Dhoubhadel BG, Hayashi Y, Domai FM, Bhattarai S, Ariyoshi K, Pandey BD., 2023. A major dengue epidemic in 2022 in Nepal: need of an efficient early-warning system. Front Trop Dis 4: 1217939

21. Auchincloss AH, Gebreab SY, Mair C, Diez Roux AV., 2012. A Review of Spatial Methods in Epidemiology, 2000–2010. Annu Rev Public Health 33: 107–122

22. Graham AJ, Atkinson PM, Danson FM., 2004. Spatial analysis for epidemiology. Acta Trop 91: 219–225

23. Oliveira MA de, Ribeiro H, Castillo-Salgado C., 2013. Geospatial analysis applied to epidemiological studies of dengue: a systematic review. Rev Bras Epidemiol Braz J Epidemiol 16: 907–917

24. Lippi CA, Stewart-Ibarra AM, Romero M, Lowe R, Mahon R, Van Meerbeeck CJ, Rollock L, Gittens-St Hilaire M, Trotman AR, Holligan D, Kirton S, Borbor-Cordova MJ, Ryan SJ., 2020. Spatiotemporal Tools for Emerging and Endemic Disease Hotspots in Small Areas: An Analysis of Dengue and Chikungunya in Barbados, 2013-2016. Am J Trop Med Hyg 103: 149–156

25. National Statistics Office., 2023. National Population and Housing Census 2021. Thapathali, Kathmandu: National Statistics Office, Office of the Prime Minister and Council of Ministers

26. Duncan JMA, Biggs EM., 2012. Assessing the accuracy and applied use of satellite-derived precipitation estimates over Nepal. Appl Geogr 34: 626–638

27. Talchabhadel R, Karki R, Thapa BR, Maharjan M, Parajuli B., 2018. Spatio-temporal variability of extreme precipitation in Nepal. Int J Climatol 38: 4296–4313

28. ECDC., 2023. 2023_12_15 Dengue Situation Update. Kathmandu, Nepal: Epidemiology and Disease Control Division, Department of Health Services, Ministry of Health and Population

29. ECDC., 2022. Situation Update of Dengue 2022 (As of 31th Dec, 2022). Kathmandu, Nepal: Epidemiology and Disease Control Division, Department of Health Services, Ministry of Health and Population

30. Central Bureau of Statistics., 2012. National Population and Housing Census 2011. Kathmandu, Nepal: National Central Bureau of Statistics, Planning Commission Secretariat

31. ESRI., 2023. ArcGIS Pro

32. Center For International Earth Science Information Network-CIESIN-Columbia University., 2016. Documentation for Gridded Population of the World, Version 4 (GPWv4). Palisades, NY: NASA Socioeconomic Data and Applications Center (SEDAC)

33. Lloyd CT, Chamberlain H, Kerr D, Yetman G, Pistolesi L, Stevens FR, Gaughan AE, Nieves JJ, Hornby G, MacManus K, Sinha P, Bondarenko M, Sorichetta A, Tatem AJ., 2019. Global spatio-temporally harmonised datasets for producing high-resolution gridded population distribution datasets. Big Earth Data 3: 108–139

34. Luc Anselin, Ibnu Syabri, Youngihn Kho., 2006. GeoDa: An Introduction to Spatial Data Analysis. Geographical Analysis 38 (1), 5–22

35. Ikejezie J, Langley T, Lewis S, Bisanzio D, Phalkey R., 2022. The epidemiology of diphtheria in Haiti, December 2014–June 2021: A spatial modeling analysis. PLOS ONE 17: e0273398

36. Parra-Amaya M, Puerta-Yepes M, Lizarralde-Bejarano D, Arboleda-Sánchez S., 2016. Early Detection for Dengue Using Local Indicator of Spatial Association (LISA) Analysis. Diseases 4: 16

37. Anselin L., 1995. Local Indicators of Spatial Association—LISA. Geogr Anal 27: 93–115

38. Anselin L, Lozano N, Koschinsky J., 2006. Rate transformations and smoothing. Urbana 51: 61801

